# Diagnostic value of PIVKA-II + AFP and PIVKA-II AFP-L3 in hepatocellular carcinoma: a systematic review and meta-analysis

**DOI:** 10.1101/2024.11.25.24317930

**Authors:** Abdulrahman Ibn Awadh, Mariam Ajeebi, Shaikhah Alharbi, Khulud Alanazi, Ahmed Alzahrani, Abdullah Alkhenizan

## Abstract

**Background:** The most promising serum biomarkers for hepatocellular carcinoma (HCC) patients include PIVKA-II, AFP, and AFP-L3. Nevertheless, the effectiveness of combining biomarkers is still up for debate. This meta-analysis aimed to evaluate the diagnostic accuracy of PIVKA-II+AFP and PIVKA-II+AFP-L3.

**Methods:** Following a systematic search of the literature in PubMed, Medline, Web of Science, Scopus, the Cochrane Library, Embase, Google Scholar and CINAHL, thirty relevant papers were found. Pooled sensitivity, specificity, and diagnostic odds ratio (DOR) were evaluated using a random-effects model.

**Results:** The pooled sensitivity, specificity and DOR values of PIVKA-II+AFP were 0,79, 0,83, and 24,95, respectively; which were slightly lower to those of PIVKA-II+AFP-L3 (0,77, 0,88, and 29,73, respectively) except for sensitivity. Furthermore, PIVKA-II+AFP-L3 presented higher diagnostic accuracy (AUC=0,923) than did PIVKA-II+AFP (AUC=0,895). Neither threshold effects nor continent or etiology of HCC were found to be sources of heterogeneity. Interestingly, we demonstrated proof of publication bias for DOR values using Egger’s regression test (p < 0,05) and funnel plots. Sensitivity analysis proved the strong reliability of this meta-analysis.

**Conclusion:** The combined assay of PIVKA-II+AFP-L3 seemed to be more adequate than PIVKA-II+AFP for the diagnosis of HCC. Hence, diagnostic tests combining many biomarkers will be clinically significant for HCC decision-making processes.

## INTRODUCTION

Liver cancer is the leading cause of cancer-related deaths worldwide and has become a significant global public health issue. According to global cancer statistics, liver cancer accounted for 4.7% of all cancer cases and 8.3% of all cancer-related deaths in 2020 [1]. Hepatocellular carcinoma (HCC) represents 70–85% of all primary liver cancers, making it the most common form of the disease [2]. Most HCC patients are diagnosed at middle or late stages because the disease does not present clinical symptoms early on. Consequently, surgical treatment is often considered, though it may adversely affect patients’ quality of life [3]. Imaging techniques remain the primary methods for monitoring and diagnosing HCC [4]. However, serum biomarker detection should be the preferred method for quick clinical decision-making due to its non-invasive nature, ease of use, low cost, high efficiency, and high throughput. The protein induced by vitamin K absence or antagonist II (PIVKA-II), first described in 1984 [5], has been identified as a suitable serum biomarker for HCC detection, with a specificity of 90% [6]. In clinical practice, alpha-fetoprotein (AFP) is the most commonly used serological biomarker, especially for high-risk cirrhosis patients, where it is often used in combination with hepatic ultrasonography for HCC detection [7]. Recent studies have identified other notable serum biomarkers, such as the Lens culinaris-agglutinin-reactive fraction of AFP (AFP-L3), which is a specific AFP subtype that binds to the lectin Lens culinaris agglutinin [8]. Evidence suggests that AFP-L3 can predict the aggressive potential of HCC, regardless of tumor size or AFP levels in the serum [9].

Patients with HCC typically show significantly higher serum levels of these biomarkers, making them valuable diagnostic indicators. The diagnostic values of these biomarkers vary considerably, with PIVKA-II having sensitivities ranging from 48%-62% and specificities from 81%-98% [10], AFP showing sensitivities of 40%-65% and specificities of 80%-94% [11], and AFP-L3 exhibiting sensitivities from 30%-70% and specificities from 90%-92% [12]. Some studies suggest that combining assays for these biomarkers offers much higher diagnostic accuracy than individual assays [13], while other studies argue that adding more biomarkers does not significantly improve the diagnostic value of AFP alone [14]. As there is ongoing debate regarding the combination assays for HCC serum biomarkers, more comprehensive analysis panels are needed to validate the diagnostic effectiveness of different biomarkers. Therefore, the aim of this meta-analysis is to assess the diagnostic accuracy of two combined tests: PIVKA-II + AFP and PIVKA-II + AFP-L3 for HCC detection.

## METHODS

This systematic review and meta-analysis study was conducted according the Preferred Reporting Items for Systematic Reviews and Meta-Analyses (PRISMA) guidelines [15]. The protocol of this systematic review was registered on 07/03/2024 at PROSPERO (registration number: CRD42024517499)

### Search strategy

The following databases were searched from database inception until January 2024: PubMed/Medline, Web of Science, Scopus, the Cochrane Library, Embase, Google Scholar and CINAHL. The search strategy was based on the following key search terms: “hepatocellular carcinoma” OR “liver cancer” OR “hepatoma” OR “HCC” AND “Des-gamma-carboxy prothrombin” OR “DCP” OR “protein induced by vitamin K absence II” OR “PIVKA-II” AND “alpha-fetoprotein” OR “AFP” AND “Lens culinaris agglutinin-reactive fraction of α-fetoprotein” OR “Alpha-fetoprotein-L3” OR “AFP-L3”. We also manually searched the references mentioned in narrative reviews and pertinent non-systematic papers to find further relevant studies that our search approach could have overlooked. All retrieval processes were performed independently by two authors ( AI and SB)

### Inclusion and exclusion criteria

Relevant articles were initially screened based on their titles and abstracts, with duplicates removed. Studies were deemed eligible if they investigated the diagnostic accuracy of PIVKA-II + AFP and/or PIVKA-II + AFP-L3 for detecting HCC. Full-text reviews were conducted on the remaining studies to confirm their inclusion.

The inclusion criteria were as follows:

– Observational studies evaluating the diagnostic performance of PIVKA-II + AFP and/or PIVKA-II + AFP-L3 for HCC.
– Studies involving patients diagnosed with HCC based on widely accepted radiological criteria.
– Publications providing data on sensitivity and specificity.
– Use of serum as the specimen type.
– Original research articles.

Exclusion criteria included:

– Unavailability of full-text articles online.
– Publications in languages other than English.
– Comments, letters, editorials, protocols, guidelines, or review articles.
– Studies with insufficient outcome data.

### Data extraction

Two independent authors ( AI and SB) retrieved information from the eligible articles following the inclusion and exclusion criteria, and information were collected on a standardized data sheet that included: (1) study ID (name of first author, year of publication), (2) country, (3) sample size of HCCs/Controls, (4) etiology of HCC, (5) type of controls, (6) mean Age (SD) or median age (range) of HCC/Control, years, (7) male, n (%), HCC/Control, (8) cut-off value of PIVKA-II, (9) cut-off value of AFP, (10) cut-off value of AFP-L3, and (11) type of combination.

### Quality assessment of the studies

The methodologic quality of the included studies was evaluated independently, by 2 authors SB and AA using the Quality Assessment of Diagnostic Accuracy Studies-2 (QUADAS-2) tool, which includes four criteria: “patient selection”, “index test”, “reference standard”, and “flow and timing” and judge bias and applicability [16]. Each is assessed in terms of risk of bias, and the first 3 domains were assessed with respect to applicability. Each item is answered with “yes,” “no,” or “unclear.” The answer of “yes” means low risk of bias, whereas “no” or “unclear” means the opposite. Any disagreements were resolved by inviting a third author AZ to participate in the discussion.

### Statistical Analysis

This diagnostic meta-analysis was conducted on the analytical software Meta-disc 1.4 and the statistical software Comprehensive Meta-Analysis version 3 (Biostat Inc. USA) in order to analyze the pooled sensitivity, specificity, Diagnostic Odds Ratio (DOR), and the area under the curve (AUC) values with 95% confidence intervals (CIs) across studies. The data were considered statistically significant when two-sided p < 0,05. The summary receiver operating characteristic (SROC) curve was also used based on the sensitivity and specificity of each study to assess the diagnostic performance. Because of the differences in the basic features of the included articles, their diverging results may have been caused by heterogeneity or random error. Therefore, the Cochrane chi-squared test was used to evaluate heterogeneity among articles, with p < 0,05 indicating the existence of heterogeneity. To estimate the impact of heterogeneity on the meta-analysis, I^2^ value was also calculated. If p <0,05 and I^2^ >50%, heterogeneity was defined as significant. In order to explore heterogeneity, the threshold effect was assessed using the Spearman correlation coefficient. A strong positive correlation would suggest a threshold effect. Meta-regression analysis was also performed to identify potential sources of heterogeneity according to the year of publication, geographical origin of studies and etiology of HCC. Furthermore, a sensitivity analysis was conducted to evaluate the validity and robustness of the meta-analysis. Finally, Egger’s test was conducted to evaluate publication bias. This latter was further assessed by the visual inspection of the symmetry in funnel plots.

## RESULTS

### Identification of studies

The database search identified 1267 studies to be screened, of which 698 abstracts were identified as potentially eligible and retrieved for full-text review. Eligibility criteria were met by 30 articles, which were included in this systematic review and meta-analysis study. The PRISMA flowchart is shown in Figure 1.

**Figure 1.**
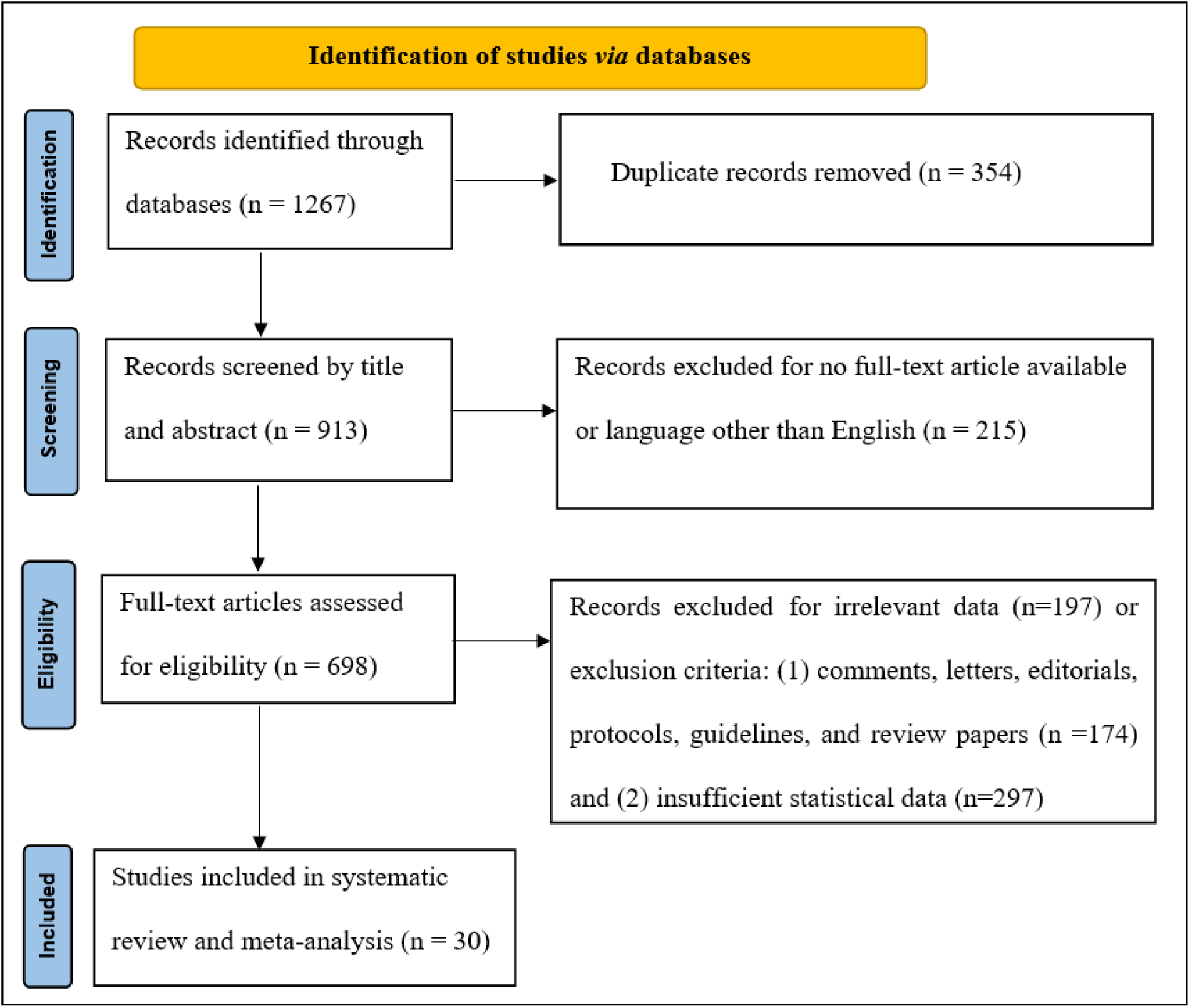
PRISMA flow diagram of the literature study process and selection.

### Characteristics of included studies

The included articles were published between 2007 and 2023 and distributed among 7 countries: China (n=15), Republic of Korea (n=5), USA (n=3), Italy (n=3), Vietnam (n=2), Germany (n=1), France (n=1). Among the 49 articles included in this systematic review and meta-analysis, 10 were cross-sectional studies and 39 were cohort studies (prospective or retrospective studies). The sample size of the included articles varied from 36 to 419 participants for HCC group and from 60 to 644 participants for control group. Twenty-nine studies reported the diagnostic effectiveness of PIVKA-II+ AFP, while ten studies compared the diagnostic effectiveness of both PIVKA-II+ AFP and PIVKA-II+ AFP-L3. However, one study described the diagnostic value of only PIVKA-II+ AFP-L3.

Characteristics of included studies are summarized in Table 1.

**Table 1.**
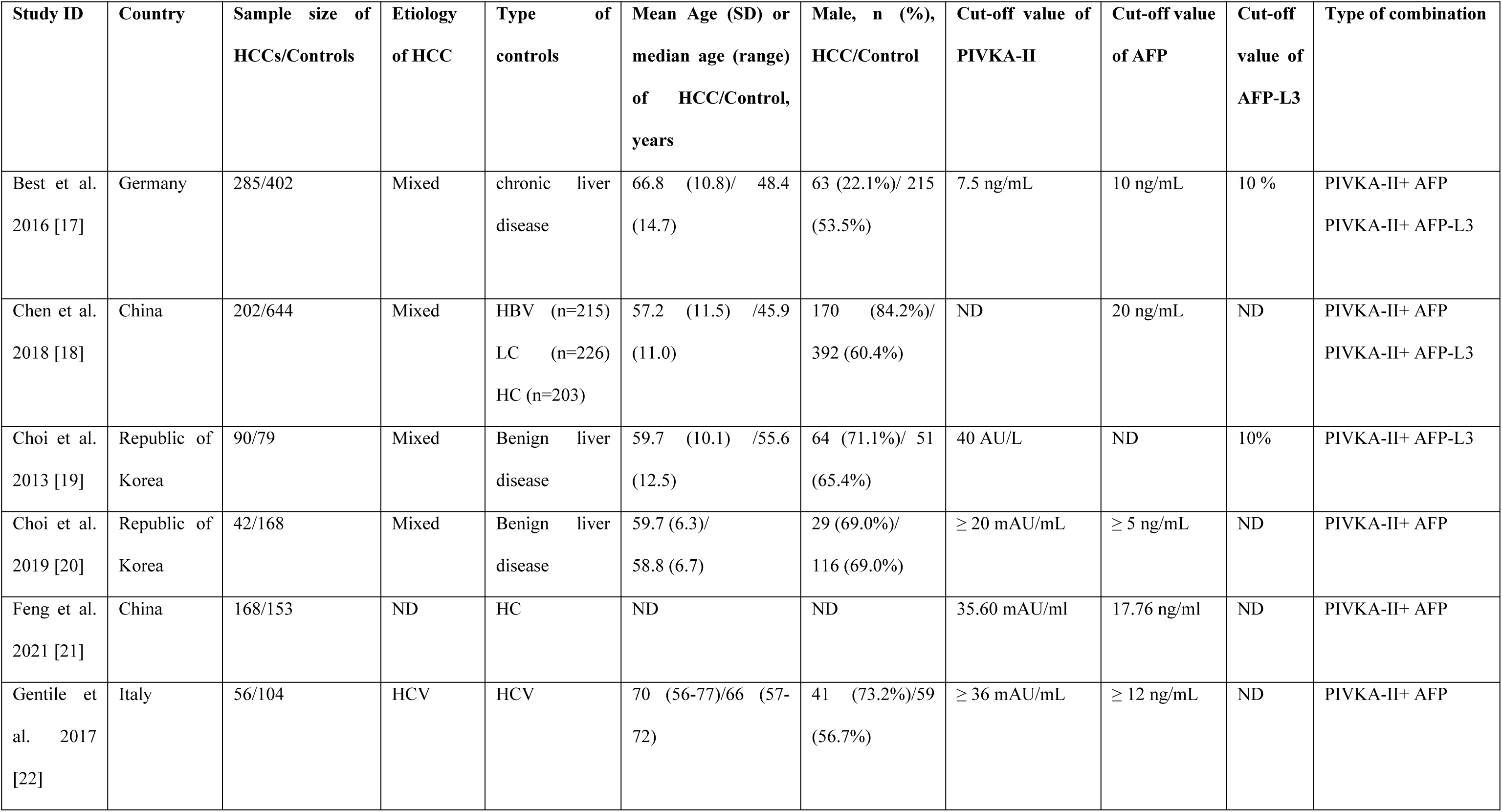
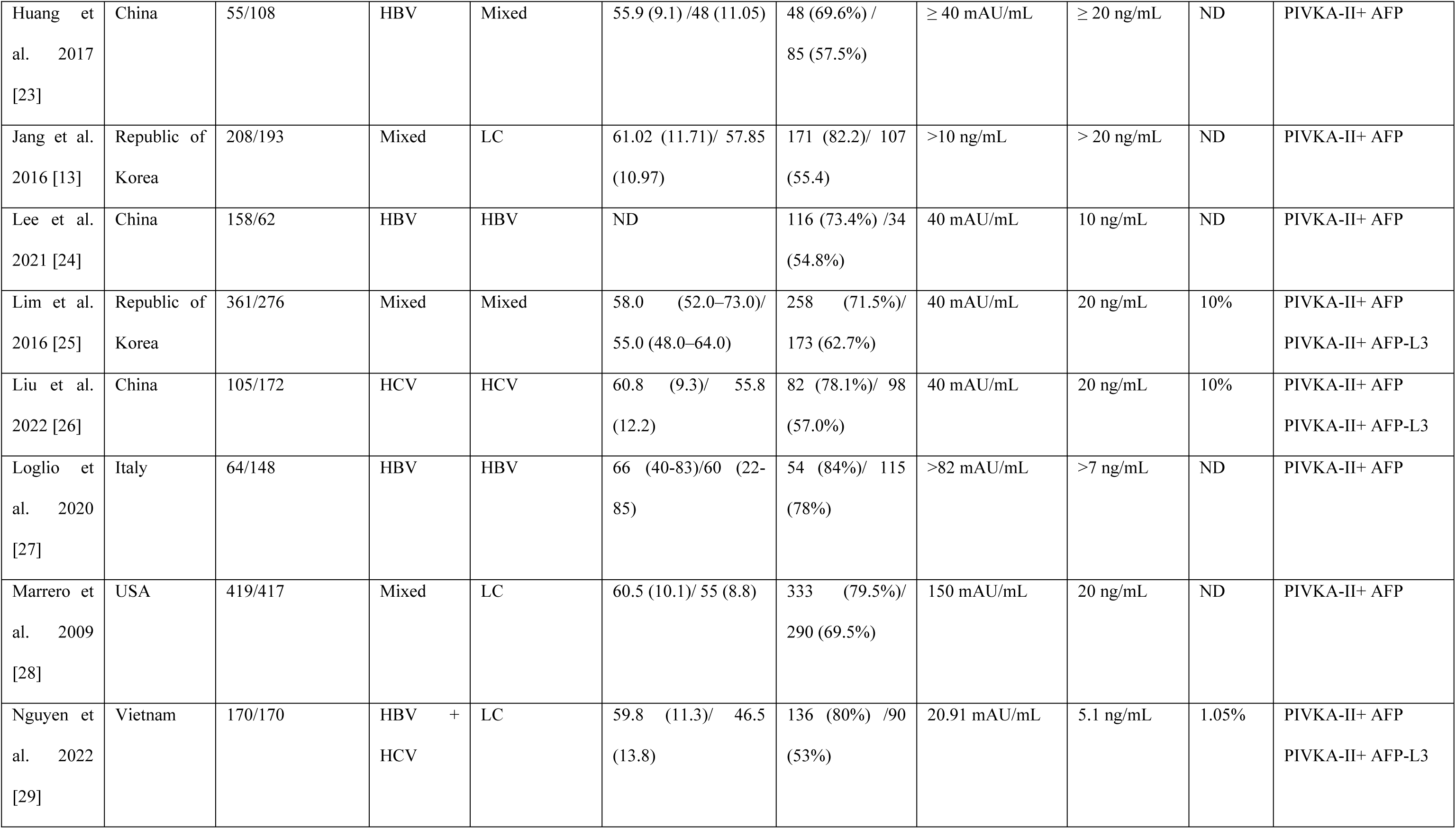
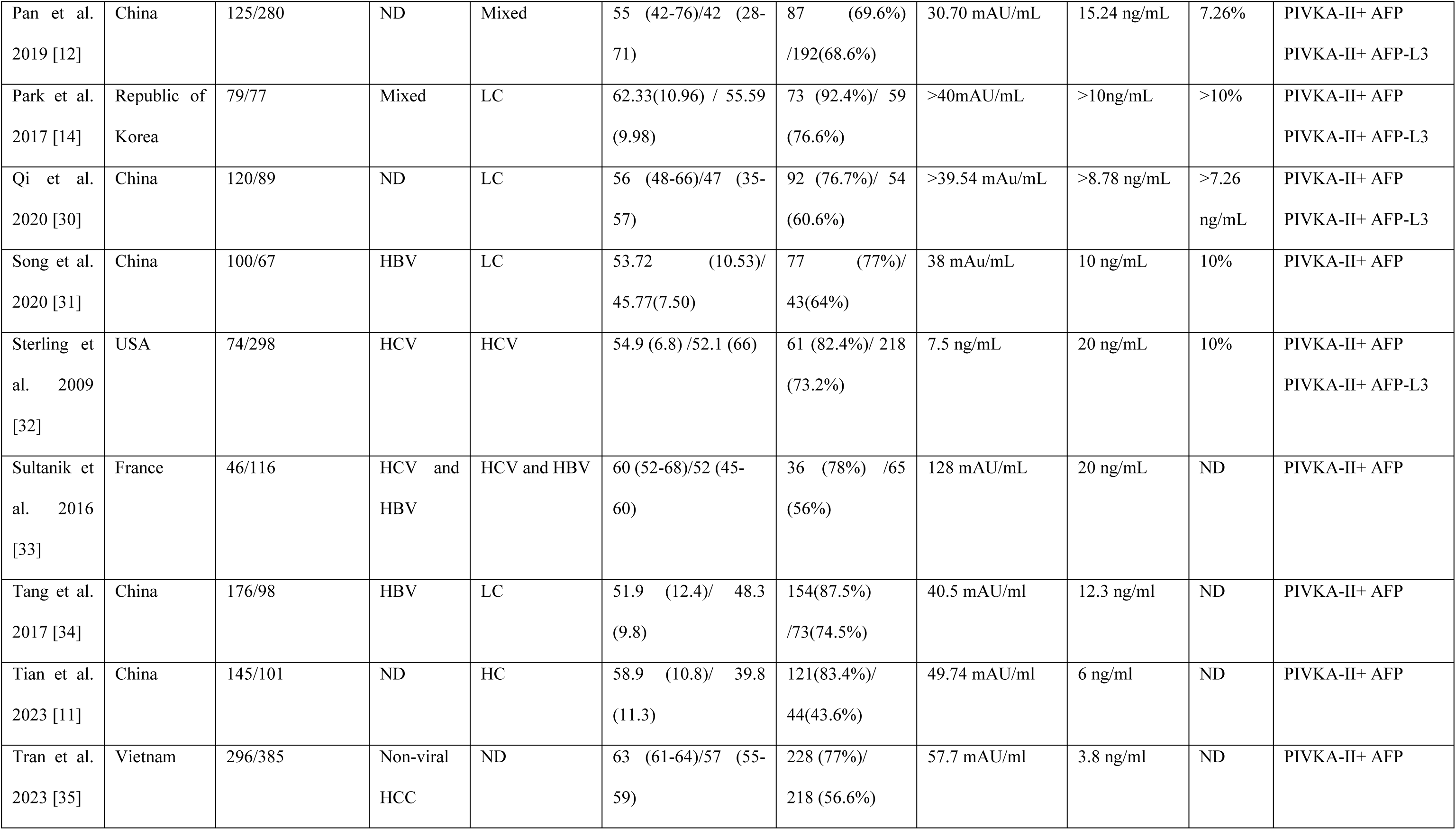
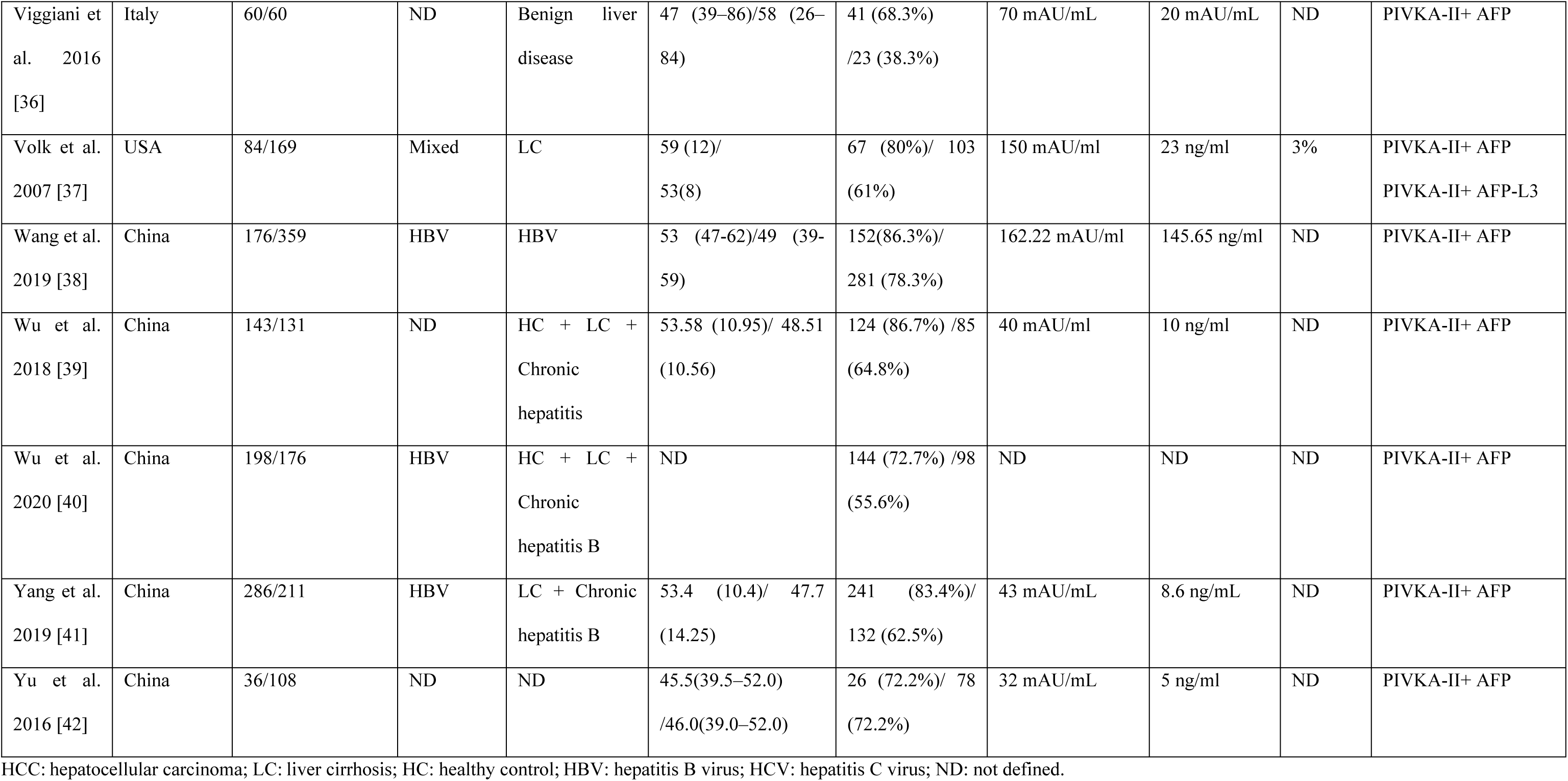
Characteristics of the studies included in the meta-analysis.

### Quality assessment of studies

The methodological quality of the 30 studies was evaluated using the QUADAS-2 tool. Regarding the patient selection domain, most studies (25 out of 30) were found to have a high risk of bias due to the absence of consecutive or random sampling. Conversely, a low risk of bias was observed in the index test domain, primarily because a preset threshold was used (1 out of 30 studies). The reference standard and flow and timing domains showed no significant risk of bias.

Additionally, there were minimal concerns about the applicability of the included studies. High applicability concerns were identified in five studies for the patient selection domain and in one study for the index test domain. Further details of the quality assessment are presented in Figure 2.

**Figure 2.**
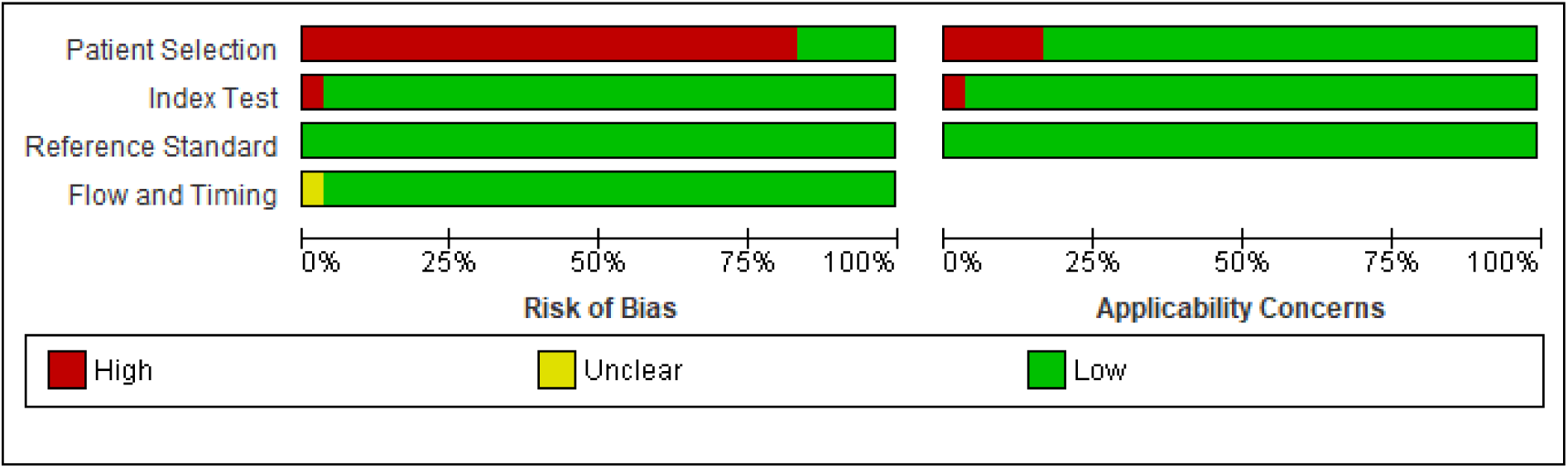
Risk of bias and applicability concerns graph: review authors’ judgments about each domain presented as percentages across included studies.

### Data analysis

#### PIVKA-II+ AFP

Twenty-nine studies reported the diagnostic accuracy of the combination of PIVKA-II+ AFP. From forest plots of pooled data, we found significant heterogeneity in sensitivity (Chi^2^=215,51, p=0,0000, I^2^= 87%, Figure 3), specificity (Chi^2^=333,74, p=0,0000, I^2^= 91,60%, Figure 4), and DOR (Chi^2^=159,40, p=0,0000, I^2^= 82,40%, Figure 5) outcomes. Consequently, the random-effect model was used to calculate the pooled estimates of these evaluation indicators.

**Figure 3.**
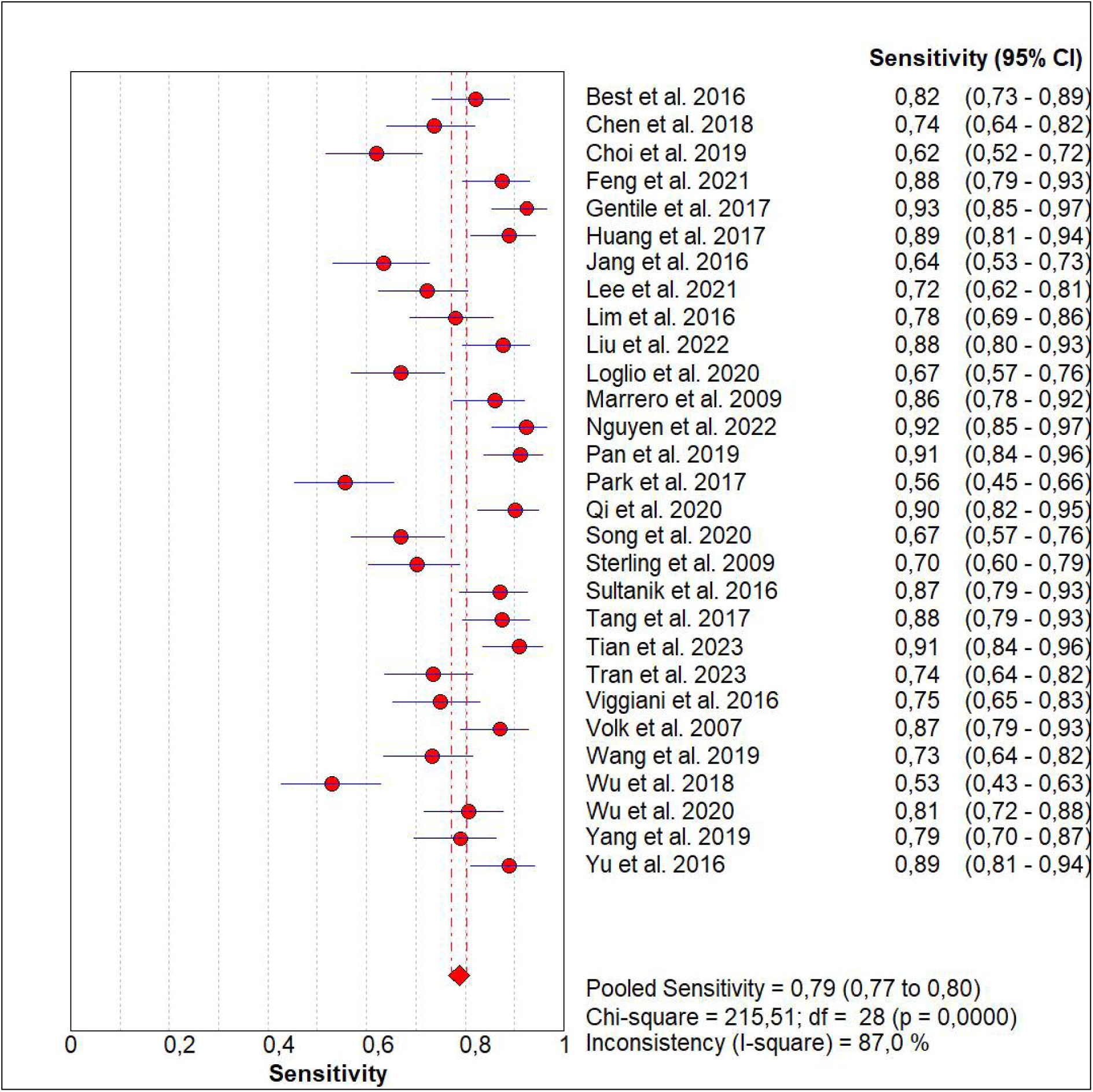
Forest plot for the sensitivity of PIVKA-II+ AFP for the diagnosis of HCC

**Figure 4.**
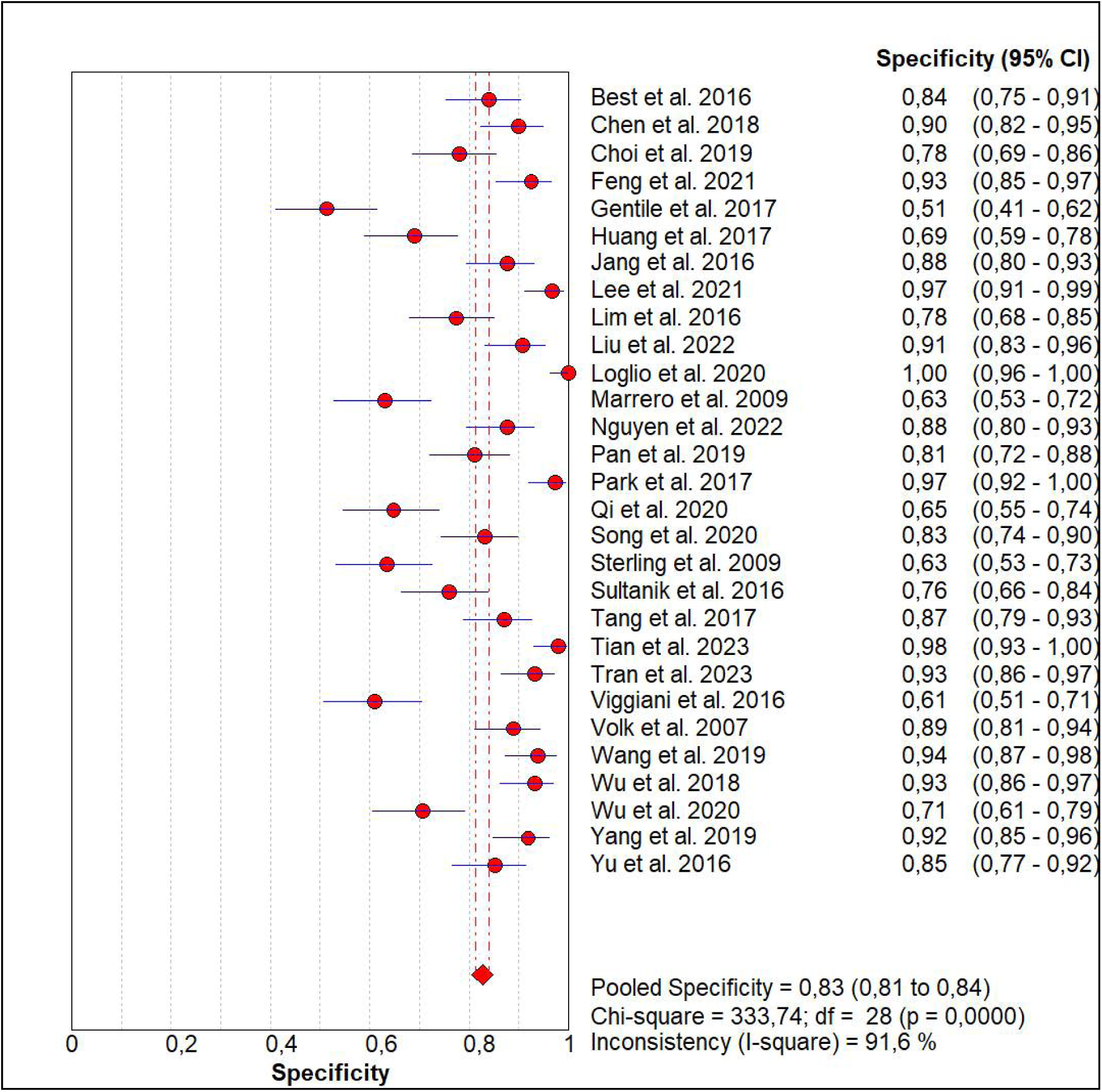
Forest plot for the specificity of PIVKA-II+ AFP for the diagnosis of HCC

**Figure 5.**
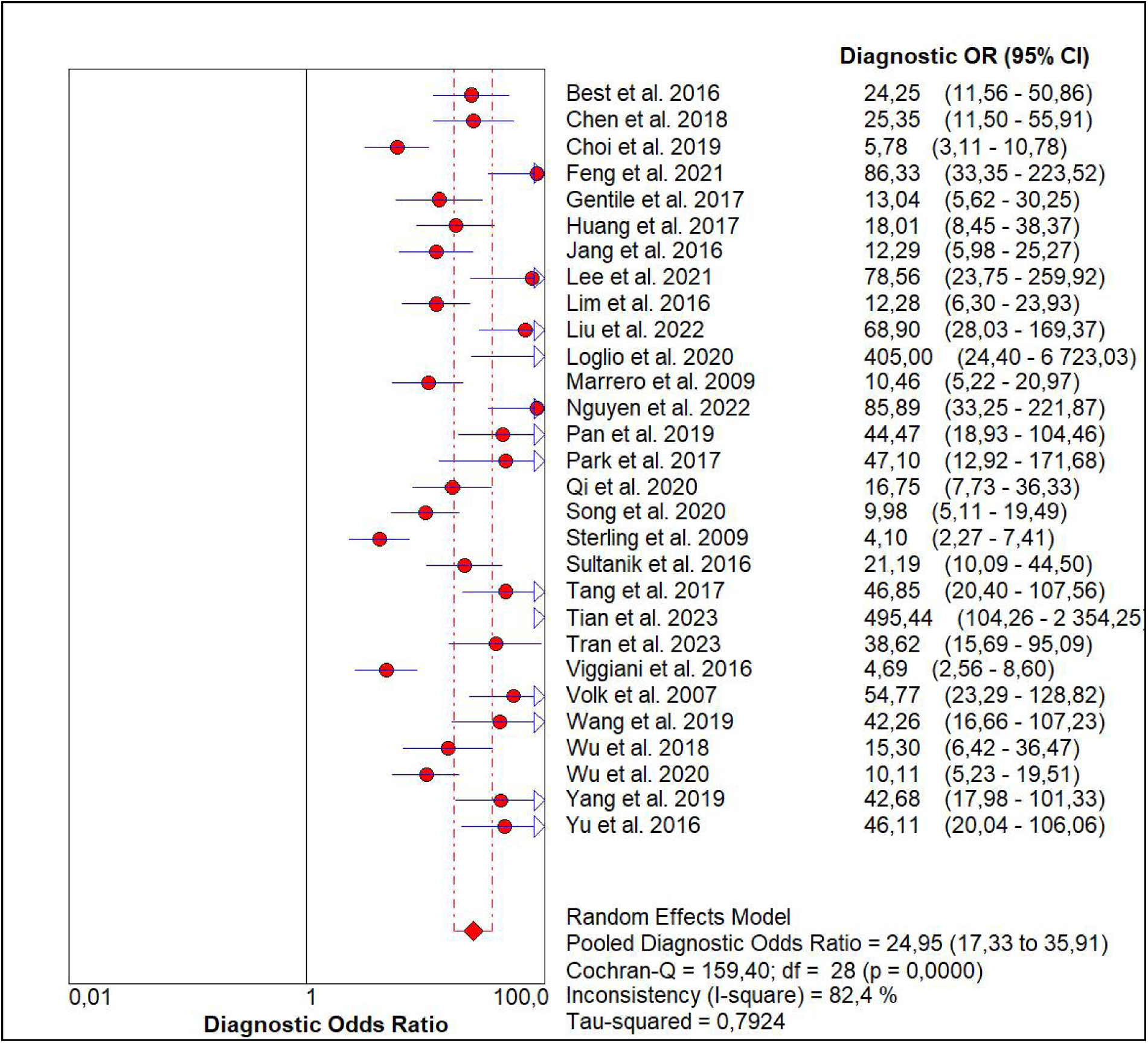
Forest plot for the DOR of PIVKA-II+ AFP for the diagnosis of HCC

The pooled estimates of PIVKA-II+ AFP for the diagnosis of HCC were 0,79 (95% CI: 0,77-0,80) for sensitivity (Figure 3), 0,83 (95% CI: 0,81-0,84) for specificity (Figure 4), and 24,95 (95% CI: 17,33-35,91) for DOR (Figure 5).

Moreover, we plotted the SROC curve to evaluate the diagnostic accuracy of PIVKA-II+ AFP (Figure 6). AUC was 0,895 (95% CI: 0,890-0,900), suggesting an outstanding diagnostic accuracy of PIVKA-II+ AFP.

**Figure 6.**
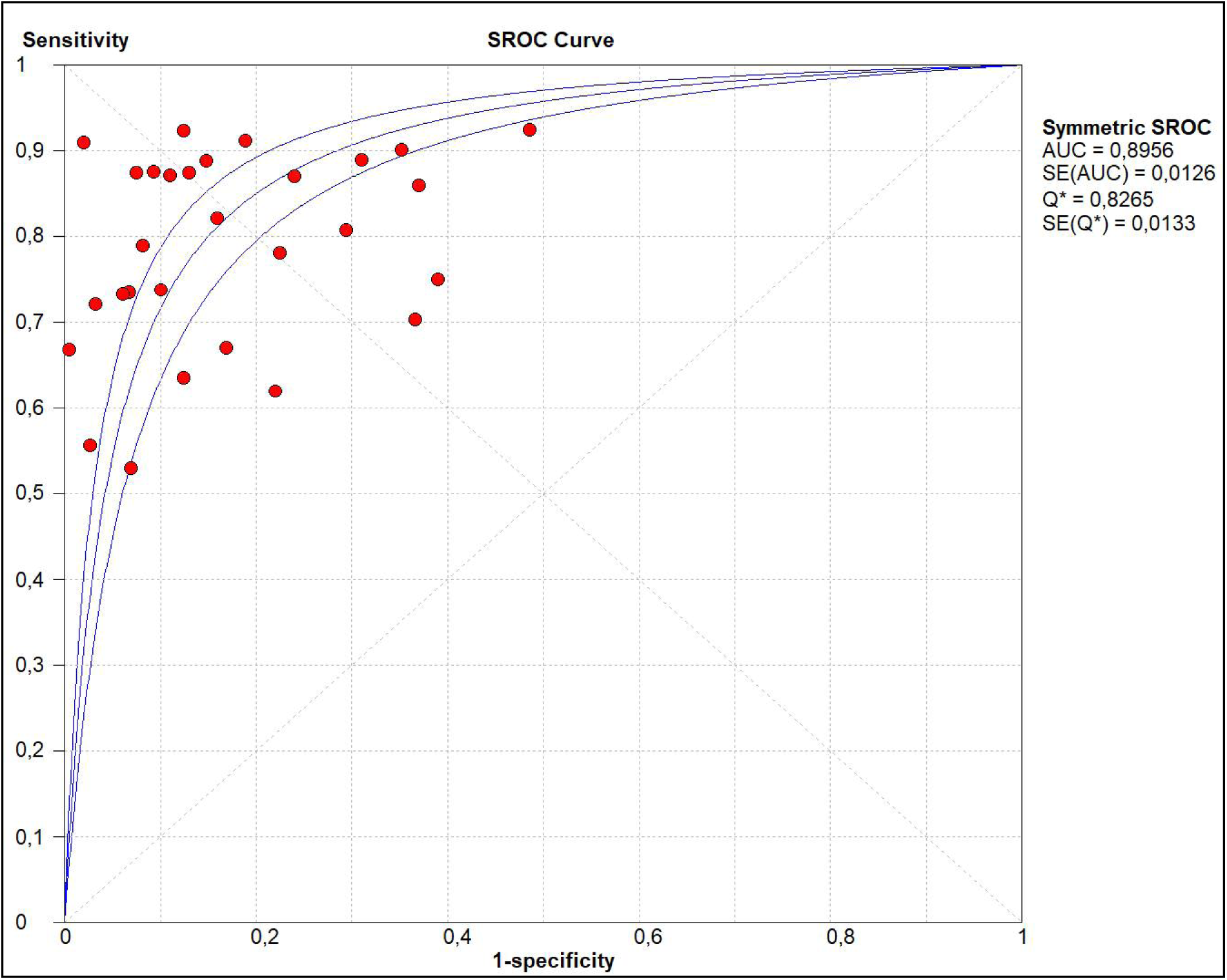
SROC curve for the diagnostic accuracy of PIVKA-II+ AFP

#### PIVKA-II+ AFP-L3

Eleven studies reported the diagnostic accuracy of the combination of PIVKA-II+ AFP-L3. From forest plots of pooled data, we found significant heterogeneity in sensitivity (Chi^2^=106,26, p=0,0000, I^2^= 90,6%, Figure 7), specificity (Chi^2^=27,8, p=0,0019, I^2^=64%, Figure 8), and DOR (Chi^2^=37,70, p=0,0000, I^2^= 73,5%, Figure 9) outcomes. Consequently, the random-effect model was used to calculate the pooled estimates of these evaluation indicators.

**Figure 7.**
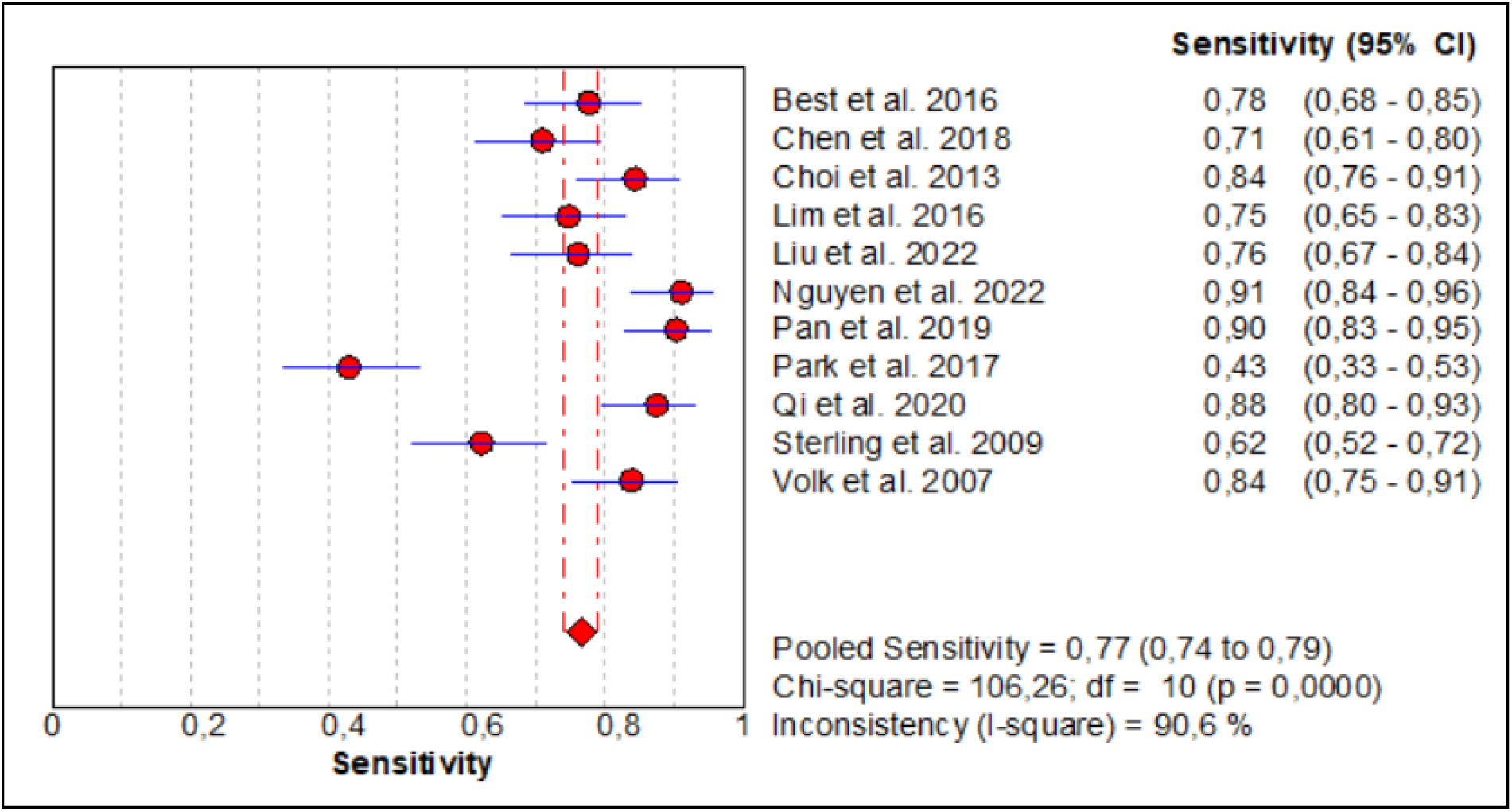
Forest plot for the sensitivity of PIVKA-II+ AFP-L3 for the diagnosis of HCC

**Figure 8.**
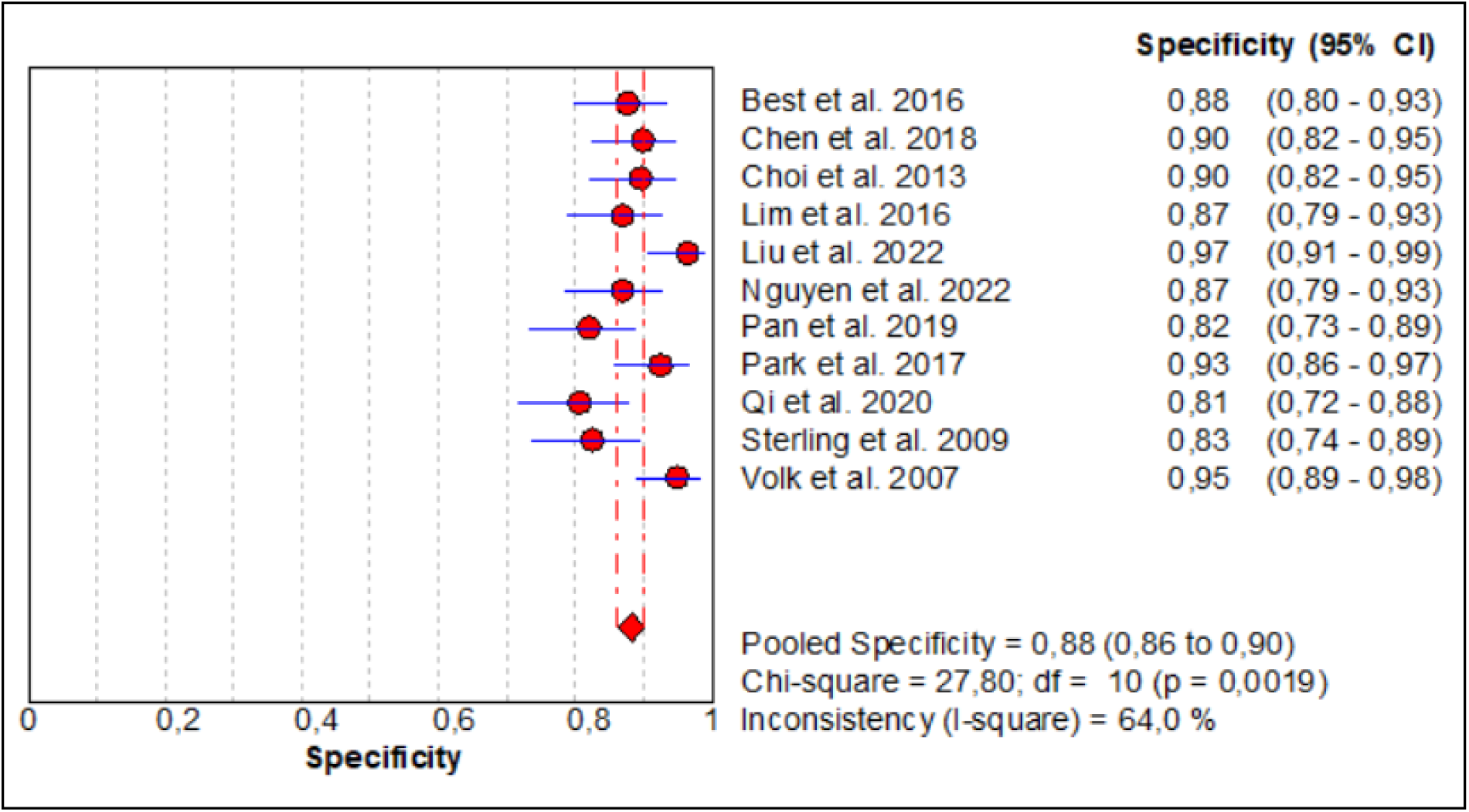
Forest plot for the specificity of PIVKA-II+ AFP-L3 for the diagnosis of HCC

**Figure 9.**
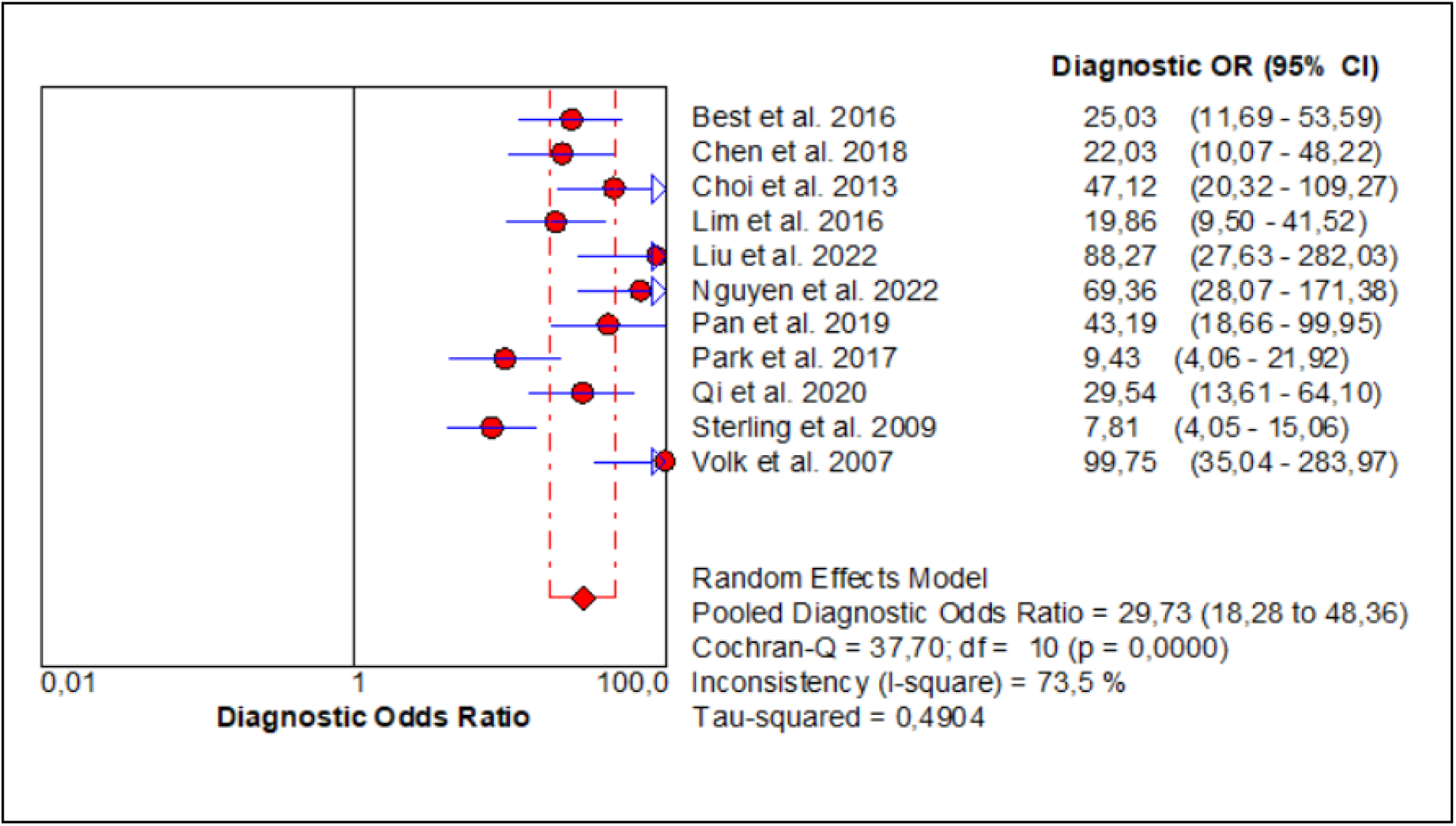
Forest plot for the DOR of PIVKA-II+ AFP-L3 for the diagnosis of HCC

The pooled estimates of PIVKA-II+ AFP for the diagnosis of HCC were 0,77 (95% CI: 0,74-0,79) for sensitivity (Figure 7), 0,88 (95% CI: 0,86-0,90) for specificity (Figure 8), and 29,73 (95% CI: 18,28-48,36) for DOR (Figure 9).

Moreover, we plotted the SROC curve to evaluate the diagnostic accuracy of PIVKA-II+ AFP-L3 (Figure 10). AUC was 0,923 (95% CI: 0,915-0,931), suggesting an outstanding diagnostic accuracy of PIVKA-II+ AFP-L3.

**Figure 10.**
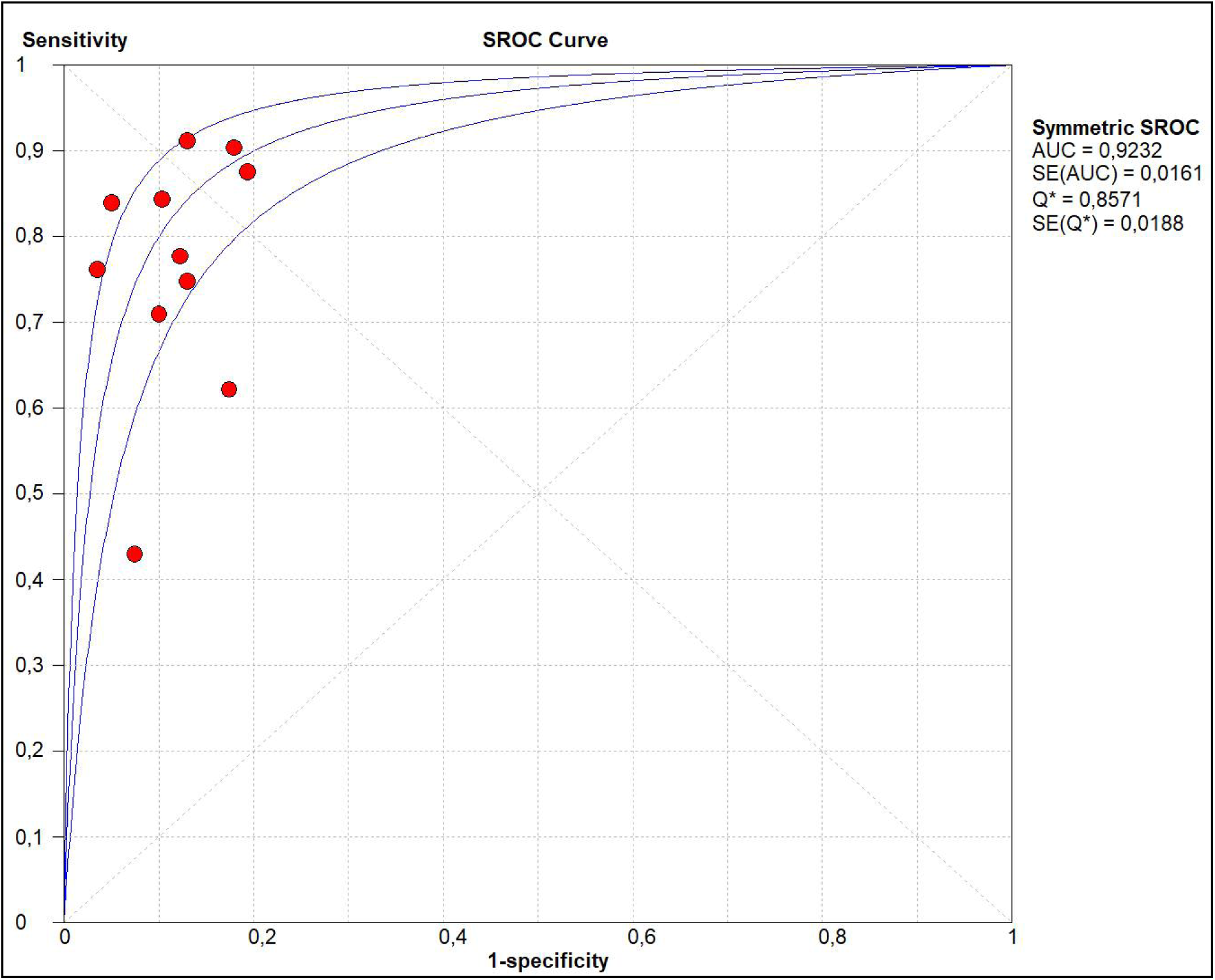
SROC curve for the diagnostic accuracy of PIVKA-II+ AFP-L3

### Threshold effects and meta-regression analysis of heterogeneity

The threshold effect is the primary source of heterogeneity in a diagnostic test. The Moses model was weighted by inverse variance, and the Spearman correlation coefficient was used to assess the threshold impact. The findings revealed that the Spearman correlation coefficient was 0,313 (p = 0,099) and 0,396 (p = 0,228) for PIVKA-II+AFP and PIVKA-II+AFP-L3, respectively. Thus, the threshold effect was not found to cause variations in the accuracy estimates among individual studies.

Apart from the threshold effect, there exist other plausible factors that could cause heterogeneity in this meta-analysis, including the etiology of HCC, the year of publication, and the geographical origin of studies. In order to examine these factors, a meta-regression analysis was carried out. Table 2 revealed that these factors did not provide any evidence for interpreting the source of heterogeneity (p>0,05). It is clear that insufficient power was provided by the current meta-regression study to comprehend the potential causes of heterogeneity.

**Table 2.**
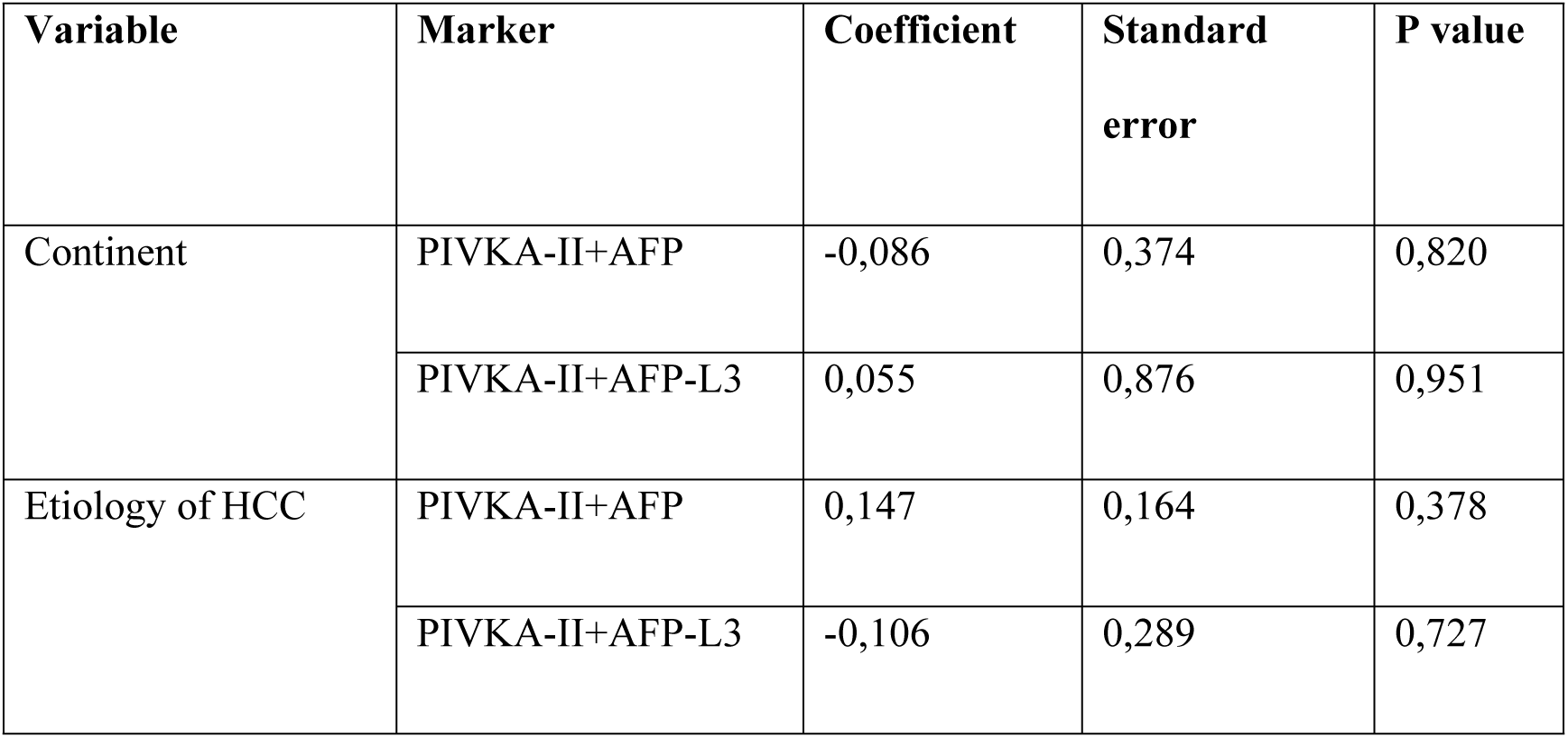
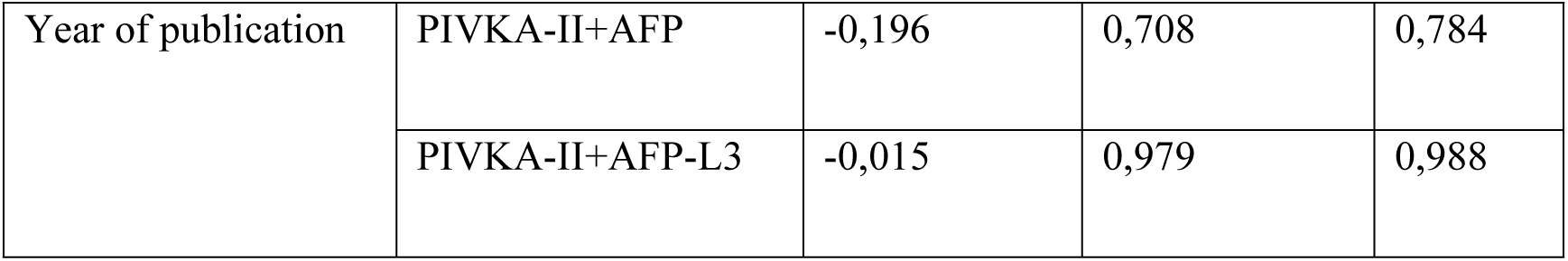
Meta-regression analysis.

### Publication bias

To assess the potential publication bias of included studies, the funnel plot symmetry test was conducted. We revealed proof of publication bias for DOR values analyzed using Egger’s regression test for PIVKA-II+AFP (p=0,0001) and PIVKA-II+AFP-L3 (p=0,0003) biomarkers. Moreover, a visual inspection of the funnel plot showed an asymmetrical funnel (Figure 11).

**Figure 11.**
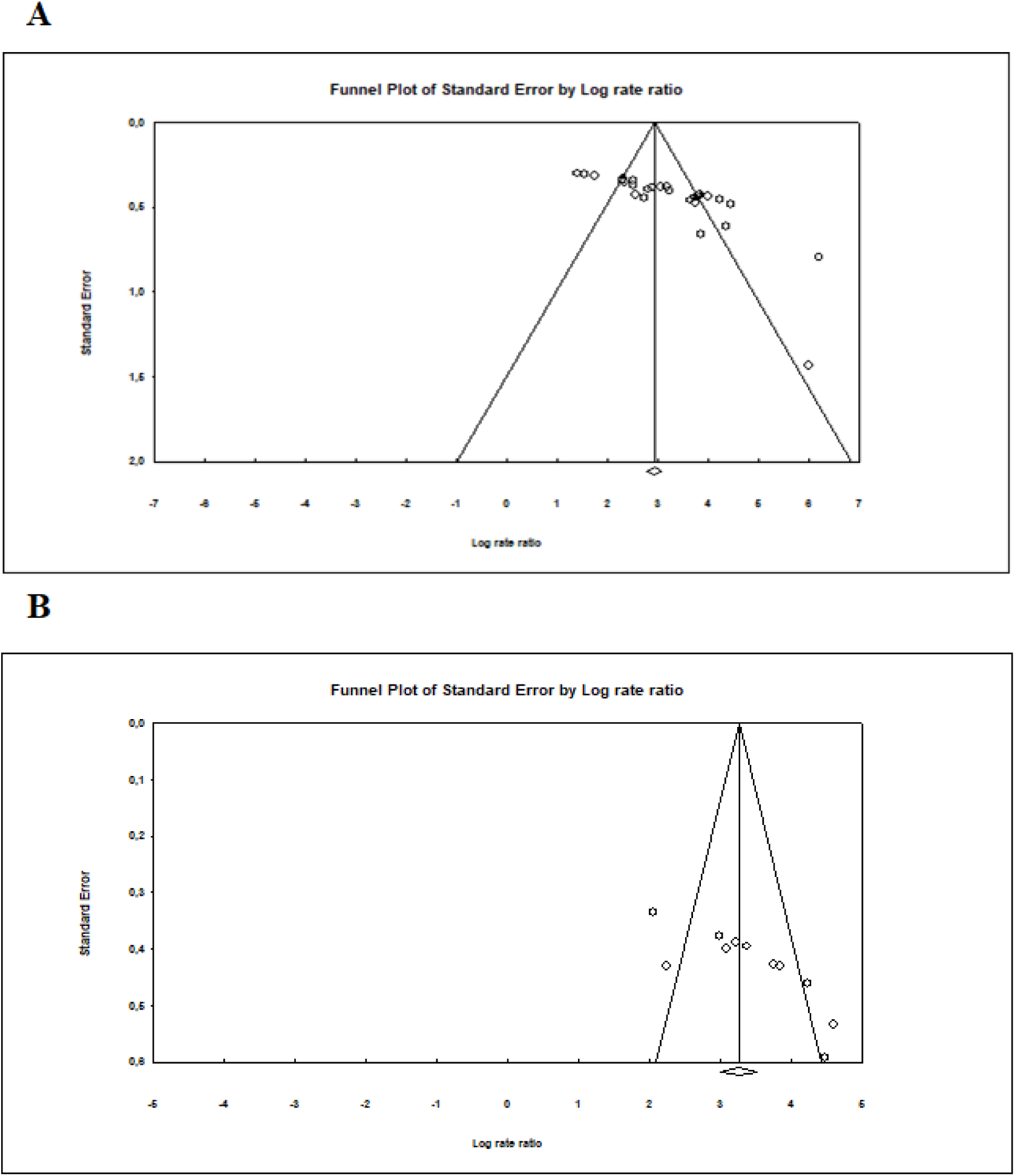
Funnel plot showing publication bias in terms of DOR values among the studies investigating (A) PIVKA-II+AFP and (B) PIVKA-II+AFP-L3.

### Sensitivity analysis

A leave-one-out sensitivity analysis was performed to assess the robustness of this meta-analysis using the pooled analysis of DOR values. The outcome did not differ markedly when a single study was omitted, which indicates that the meta-analysis had strong reliability (Table 2). Indeed, the DOR values of PIVKA-II+AFP ranged from 17,88 (95% CI: 15,38-20,80) to 20,60 (95% CI: 17,66-24,03). Similarly, the DOR values of PIVKA-II+AFP-L3 ranged from 24,09 (95% CI: 18,67-31,09) to 31,88 (95% CI: 24,39-41,65) (Table 2).

**Table 2.** Sensitivity analysis of DOR outcome.

| Study ID | PIVKA-II+AFP<br>DOR (95% CI), p | PIVKA-II+AFP-L3<br>DOR (95% CI), p |
| --- | --- | --- |
| Best et al. 2016 [17] | 18,38 (15,79-21,41),<br>p=0,000 | 26,22 (20,17-30,07),<br>p=0,000 |
| Chen et al. 2018 [18] | 18,38 (15,79-21,39),<br>p=0,000 | 26,58 (20,47-34,52),<br>p=0,000 |
| Choi et al. 2013 [19] | NA | 24,67 (19,03-31,96),<br>p=0,000 |
| Choi et al. 2019 [20] | 19,96 (17,12-23,28),<br>p=0,000 | NA |
| Feng et al. 2021 [21] | 17,89 (15,38-20,80),<br>p=0,000 | NA |
| Gentile et al. 2017 [22] | 18,81 (16,16-21,88),<br>p=0,000 | NA |
| Huang et al. 2017 [23] | 18,61 (15,99-21,67),<br>p=0,000 | NA |
| Jang et al. 2016 [13] | 18,94 (16,26-22,06),<br>p=0,000 | NA |
| Lee et al. 2021 [24] | 18,17 (15,64-21,12),<br>p=0,000 | NA |
| Lim et al. 2016 [25] | 19,00 (16,30-22,14),<br>p=0,000 | 27,01 (20,76-35,14),<br>p=0,000 |
| Liu et al. 2022 [26] | 17,92 (15,40-20,84),<br>p=0,000 | 24,62 (19,10-31,72),<br>p=0,000 |
| Loglio et al. 2020 [27] | 18,43 (15,87-21,40),<br>p=0,000 | NA |
| Marrero et al. 2009 [28] | 19,11 (16,41-22,27),<br>p=0,000 | NA |
| Nguyen et al. 2022 [29] | 17,88 (15,38-20,80),<br>p=0,000 | 24,10 (18,63-31,18),<br>p=0,000 |
| Pan et al. 2019 [12] | 18,09 (15,55-21,33),<br>p=0,000 | 24,86 (19,18-32,22),<br>p=0,000 |
| Park et al. 2017 [14] | 18,36 (15,80-21,33),<br>p=0,000 | 28,72 (22,16-37,22),<br>p=0,000 |
| Qi et al. 2020 [30] | 18,67 (16,03-21,73),<br>p=0,000 | 25,72 (19,80-33,41),<br>p=0,000 |
| Song et al. 2020 [31] | 19,21 (16,48-22,38),<br>p=0,000 | NA |
| Sterling et al. 2009 [32] | 20,60 (17,66-24,03),<br>p=0,000 | 31,88 (24,39-41,65),<br>p=0,000 |
| Sultanik et al. 2016 [33] | 18,49 (15,88-21,53),<br>p=0,000 | NA |
| Tang et al. 2017 [34] | 18,03 (15,49-20,98),<br>p=0,000 | NA |
| Tian et al. 2023 [11] | 18,04 (15,53-20,95),<br>p=0,000 | NA |
| Tran et al. 2023 [35] | 18,21 (15,66-21,18),<br>p=0,000 | NA |
| Viggiani et al. 2016 [36] | 20,32 (17,42-23,70),<br>p=0,000 | NA |
| Volk et al. 2007 [37] | 17,97 (15,45-20,91),<br>p=0,000 | 24,09 (18,67-31,09),<br>p=0,000 |
| Wang et al. 2019 [38] | 18,19 (15,64-21,16),<br>p=0,000 | NA |
| Wu et al. 2018 [39] | 18,70 (16,07-21,76),<br>p=0,000 | NA |
| Wu et al. 2020 [40] | 19,21 (16,49-22,39),<br>p=0,000 | NA |
| Yang et al. 2019 [41] | 18,12 (15,58-21,09),<br>p=0,000 | NA |
| Yu et al. 2016 [42] | 18,04 (15,50-20,99),<br>p=0,000 | NA |

NA: Not applicable.

## DISCUSSION

Hepatocellular carcinoma (HCC) is a major global public health challenge. Early detection is crucial for effective treatment, improved patient outcomes, and prolonged survival. While serum immunological assays for protein biomarkers remain practical for HCC follow-up, recent studies exploring molecular biomarkers, such as non-coding RNA, have aimed to identify gene barcodes for HCC diagnosis [43]. To enhance the diagnostic performance of protein biomarkers, efforts should focus on combining individual assays into panels.

In this meta-analysis, we evaluated the diagnostic accuracy of double biomarker combinations for HCC diagnosis, addressing the limitations of single biomarker assays and the varying conclusions of prior studies [13,14,40]. To our knowledge, this is the first meta-analysis exclusively examining the combined panels of PIVKA-II + AFP and PIVKA-II + AFP-L3. Sensitivity, specificity, diagnostic odds ratio (DOR), and area under the curve (AUC) were used as outcome measures to assess the diagnostic performance. Our goal was to achieve an optimal balance between improved diagnostic accuracy and the trade-off between sensitivity and specificity, as increased sensitivity often leads to reduced specificity [13].

The DOR, a single metric combining sensitivity and specificity, is defined as the ratio of the likelihood that a person with the disease will test positive to the likelihood that a person without the disease will test negative [44]. DOR values range from 0 to infinity, with higher values indicating better diagnostic performance. Among the combinations analyzed, the PIVKA-II + AFP-L3 panel demonstrated superior performance, with a DOR of 29.73 compared to 24.95 for the PIVKA-II + AFP panel.

The SROC curve and AUC are essential tools for evaluating diagnostic accuracy in meta-analyses. The SROC curve provides a comprehensive assessment beyond sensitivity and specificity alone [45,46]. The AUC, an index derived from the SROC curve, ranges from 0 (a test with no diagnostic value) to 1 (a perfect test). The AUC findings in this study aligned with the DOR values, reinforcing the conclusion that double biomarker assays offer greater clinical benefits compared to single biomarker assays. In previous meta-analyses, the AUCs of PIVKA-II, AFP, and AFP-L3 were 0.797, 0.835, and 0.710, respectively [48], which were lower than the AUCs of the combined biomarkers identified in this study.

High heterogeneity was observed in both biomarker combinations, as indicated by elevated I² values. A meta-regression analysis was conducted to explore potential sources of heterogeneity, excluding threshold effects. However, factors such as continent, HCC etiology, and publication year did not account for the observed heterogeneity. Consequently, the exact sources of heterogeneity remain unexplained.

Sensitivity analyses, performed by removing each study in turn, showed stable DOR outcomes for both combinations, underscoring the reliability of this meta-analysis. However, publication bias was identified in both panels, highlighting the need for further research to validate these findings.

This meta-analysis offers several advantages over previous studies [47,49–51]. First, it evaluates the clinical utility of combining assays from three distinct biomarkers instead of relying on a single biomarker for HCC diagnosis. Second, it includes a comprehensive review of recent literature, providing updated insights into combined biomarker panels. Third, it presents a novel perspective on the clinical application of these biomarkers and addresses concerns about the efficacy of combining them.

Despite these strengths, some limitations exist. Promising biomarkers such as GP73, DKK-1, and GPC3 were not included, though their potential clinical value warrants further exploration. Additionally, standardization of assay interpretation and evaluation is urgently needed, as variations in laboratory methodologies reduce comparability and uniformity. Finally, factors such as HCC etiology, clinical stage, and control group heterogeneity should have been more thoroughly considered in this analysis.

Further research is necessary to investigate the diagnostic accuracy of combined biomarker panels, including additional biomarkers, and to establish standardized protocols for their clinical application.

## CONCLUSION

The results of this meta-analysis showed that the double biomarker assay with the PIVKA-II and AFP-L3 panel exhibited significantly higher diagnostic accuracy for HCC than did PIVKA-II and AFP panel. At this point, new approaches are desperately needed to standardize different clinical studies in terms of comparability, timeliness, and reproducibility for the detection of HCC biomarkers. Moreover, additional biomarkers should be combined with PIVKA-II and AFP-L3 for comprehensive diagnosis of HCC.

## Data Availability Statement

All data analyzed during this meta-analysis are included in the reference section of this article and its supplementary information files.

## Declarations

## Ethical Approval

Not applicable

## Funding

No Funding.

## Notes

We have No conflicts of interest.

### Competing Interest Statement

The authors have declared no competing interest.

### Clinical Trial

NA

### Funding Statement

The author(s) received no specific funding for this work.

### Author Declarations

Its meta-analysis study.

## References

[1] Sung H, Ferlay J, Siegel RL, Laversanne M, Soerjomataram I, Jemal A, et al. Global Cancer Statistics 2020: GLOBOCAN Estimates of Incidence and Mortality Worldwide for 36 Cancers in 185 Countries. CA A Cancer J Clinicians 2021;71:209–49. 10.3322/caac.21660.

[2] Delgado TC, Barbier-Torres L, Zubiete-Franco I, Lopitz-Otsoa F, Varela-Rey M, Fernández-Ramos D, et al. Neddylation, a novel paradigm in liver cancer. Transl Gastroenterol Hepatol 2018;3:37–37. 10.21037/tgh.2018.06.05.

[3] Tang A, Hallouch O, Chernyak V, Kamaya A, Sirlin CB. Epidemiology of hepatocellular carcinoma: target population for surveillance and diagnosis. Abdom Radiol 2018;43:13–25. 10.1007/s00261-017-1209-1.

[4] Ayuso C, Rimola J, Vilana R, Burrel M, Darnell A, García-Criado Á, et al. Diagnosis and staging of hepatocellular carcinoma (HCC): current guidelines. European Journal of Radiology 2018;101:72–81. 10.1016/j.ejrad.2018.01.025.

[5] Liebman HA, Furie BC, Tong MJ, Blanchard RA, Lo K-J, Lee S-D, et al. Des-γ-Carboxy (Abnormal) Prothrombin as a Serum Marker of Primary Hepatocellular Carcinoma. N Engl J Med 1984;310:1427–31. 10.1056/NEJM198405313102204.

[6] Hadi H, Wan Shuaib WMA, Raja Ali RA, Othman H. Utility of PIVKA-II and AFP in Differentiating Hepatocellular Carcinoma from Non-Malignant High-Risk Patients. Medicina 2022;58:1015. 10.3390/medicina58081015.

[7] Stefaniuk P, Cianciara J, Wiercinska-Drapalo A. Present and future possibilities for early diagnosis of hepatocellular carcinoma. WJG 2010;16:418. 10.3748/wjg.v16.i4.418.

[8] Luo P, Wu S, Yu Y, Ming X, Li S, Zuo X, et al. Current Status and Perspective Biomarkers in AFP Negative HCC: Towards Screening for and Diagnosing Hepatocellular Carcinoma at an Earlier Stage. Pathol Oncol Res 2020;26:599–603. 10.1007/s12253-019-00585-5.

[9] Oka H, Saito A, Ito K, Kumada T, Satomura S, Kasugai H, et al. Multicenter prospective analysis of newly diagnosed hepatocellular carcinoma with respect to the percentage of *Lens culinaris* agglutinin-reactive α-fetoprotein ^1^. J of Gastro and Hepatol 2001;16:1378–83. 10.1046/j.1440-1746.2001.02643.x.

[10] Abd El Gawad IA, Mossallam GI, Radwan NH, Elzawahry HM, Elhifnawy NM. Comparing Prothrombin induced by vitamin K absence-II (PIVKA-II) with the oncofetal proteins Glypican-3, Alpha feto protein and Carcinoembryonic antigen in diagnosing hepatocellular carcinoma among Egyptian patients. Journal of the Egyptian National Cancer Institute 2014;26:79–85. 10.1016/j.jnci.2014.01.001.

[11] Tian S, Chen Y, Zhang Y, Xu X. Clinical value of serum AFP and PIVKA-II for diagnosis, treatment and prognosis of hepatocellular carcinoma. Clinical Laboratory Analysis 2023;37:e24823. 10.1002/jcla.24823.

[12] Pan Y, Dai J, Hua X, Chen J, Chen Y, Liao Y. THE VALUE OF COMBINED DETECTION OF PIVKA-II, AFP AND AFP-L3 IN THE DIAGNOSIS OF HEPATOCELLULAR CARCINOMA. Acta Medica Mediterranea 2019:2793–7. 10.19193/0393-6384_2019_5_439.

[13] Jang ES, Jeong S-H, Kim J-W, Choi YS, Leissner P, Brechot C. Diagnostic Performance of Alpha-Fetoprotein, Protein Induced by Vitamin K Absence, Osteopontin, Dickkopf-1 and Its Combinations for Hepatocellular Carcinoma. PLoS ONE 2016;11:e0151069. 10.1371/journal.pone.0151069.

[14] Park SJ, Jang JY, Jeong SW, Cho YK, Lee SH, Kim SG, et al. Usefulness of AFP, AFP-L3, and PIVKA-II, and their combinations in diagnosing hepatocellular carcinoma. Medicine 2017;96:e5811. 10.1097/MD.0000000000005811.

[15] Liberati A, Altman DG, Tetzlaff J, Mulrow C, Gøtzsche PC, Ioannidis JPA, et al. The PRISMA statement for reporting systematic reviews and meta-analyses of studies that evaluate health care interventions: explanation and elaboration. Journal of Clinical Epidemiology 2009;62:e1–34. 10.1016/j.jclinepi.2009.06.006.

[16] Whiting PF, Rutjes AWS, Westwood ME, Mallett S, Deeks JJ, Reitsma JB, et al. QUADAS-2: a revised tool for the quality assessment of diagnostic accuracy studies. Ann Intern Med 2011;155:529–36. 10.7326/0003-4819-155-8-201110180-00009.

[17] Best J, Bilgi H, Heider D, Schotten C, Manka P, Bedreli S, et al. The GALAD scoring algorithm based on AFP, AFP-L3, and DCP significantly improves detection of BCLC early stage hepatocellular carcinoma. Z Gastroenterol 2016;54:1296–305. 10.1055/s-0042-119529.

[18] Chen H, Zhang Y, Li S, Li N, Chen Y, Zhang B, et al. Direct comparison of five serum biomarkers in early diagnosis of hepatocellular carcinoma. CMAR 2018;Volume 10:1947–58. 10.2147/CMAR.S167036.

[19] Choi JY, Jung SW, Kim HY, Kim M, Kim Y, Kim DG, et al. Diagnostic value of AFP-L3 and PIVKA-II in hepatocellular carcinoma according to total-AFP. WJG 2013;19:339. 10.3748/wjg.v19.i3.339.

[20] Choi J, Kim G, Han S, Lee W, Chun S, Lim Y. Longitudinal Assessment of Three Serum Biomarkers to Detect Very Early-Stage Hepatocellular Carcinoma. Hepatology 2019;69:1983–94. 10.1002/hep.30233.

[21] Feng H, Li B, Li Z, Wei Q, Ren L. PIVKA-II serves as a potential biomarker that complements AFP for the diagnosis of hepatocellular carcinoma. BMC Cancer 2021;21:401. 10.1186/s12885-021-08138-3.

[22] Gentile I, Buonomo AR, Scotto R, Zappulo E, Carriero C, Piccirillo M, et al. Diagnostic Accuracy of PIVKA-II, Alpha-Fetoprotein and a Combination of both in Diagnosis of Hepatocellular Carcinoma in Patients Affected by Chronic HCV Infection. In Vivo 2017;31:695–700. 10.21873/invivo.11115.

[23] Huang S, Jiang F, Wang Y, Yu Y, Ren S, Wang X, et al. Diagnostic performance of tumor markers AFP and PIVKA-II in Chinese hepatocellular carcinoma patients. Tumour Biol 2017;39:101042831770576. 10.1177/1010428317705763.

[24] Lee Q, Yu X, Yu W. The value of PIVKA-Ⅱ versus AFP for the diagnosis and detection of postoperative changes in hepatocellular carcinoma. Journal of Interventional Medicine 2021;4:77–81. 10.1016/j.jimed.2021.02.004.

[25] Lim TS, Kim DY, Han K-H, Kim H-S, Shin SH, Jung KS, et al. Combined use of AFP, PIVKA-II, and AFP-L3 as tumor markers enhances diagnostic accuracy for hepatocellular carcinoma in cirrhotic patients. Scandinavian Journal of Gastroenterology 2016;51:344–53. 10.3109/00365521.2015.1082190.

[26] Liu S, Sun L, Yao L, Zhu H, Diao Y, Wang M, et al. Diagnostic Performance of AFP, AFP-L3, or PIVKA-II for Hepatitis C Virus-Associated Hepatocellular Carcinoma: A Multicenter Analysis. JCM 2022;11:5075. 10.3390/jcm11175075.

[27] Loglio A, Iavarone M, Facchetti F, Di Paolo D, Perbellini R, Lunghi G, et al. The combination of PIVKA-II and AFP improves the detection accuracy for HCC in HBV caucasian cirrhotics on long-term oral therapy. Liver International 2020;40:1987–96. 10.1111/liv.14475.

[28] Marrero JA, Feng Z, Wang Y, Nguyen MH, Befeler AS, Roberts LR, et al. α-Fetoprotein, Des-γ Carboxyprothrombin, and Lectin-Bound α-Fetoprotein in Early Hepatocellular Carcinoma. Gastroenterology 2009;137:110–8. 10.1053/j.gastro.2009.04.005.

[29] Nguyen HB, Le X-TT, Nguyen HH, Vo TT, Le MK, Nguyen NT, et al. Diagnostic Value of hTERT mRNA and in Combination With AFP, AFP-L3%, Des-γ-carboxyprothrombin for Screening of Hepatocellular Carcinoma in Liver Cirrhosis Patients HBV or HCV-Related. Cancer Inform 2022;21:117693512211007. 10.1177/11769351221100730.

[30] Qi F, Zhou A, Yan L, Yuan X, Wang D, Chang R, et al. The diagnostic value of PIVKA-II, AFP, AFP-L3, CEA, and their combinations in primary and metastatic hepatocellular carcinoma. Clinical Laboratory Analysis 2020;34:e23158. 10.1002/jcla.23158.

[31] Song T, Wang L, Su B, Zeng W, Jiang T, Zhang T, et al. Diagnostic value of alpha-fetoprotein, *Lens culinaris* agglutinin-reactive alpha-fetoprotein, and des-gamma-carboxyprothrombin in hepatitis B virus-related hepatocellular carcinoma. J Int Med Res 2020;48:030006051988927. 10.1177/0300060519889270.

[32] Sterling RK, Jeffers L, Gordon F, Venook AP, Reddy KR, Satomura S, et al. Utility of Lens culinaris Agglutinin-Reactive Fraction of α-Fetoprotein and Des-Gamma-Carboxy Prothrombin, Alone or in Combination, as Biomarkers for Hepatocellular Carcinoma. Clinical Gastroenterology and Hepatology 2009;7:104–13. 10.1016/j.cgh.2008.08.041.

[33] Sultanik P, Ginguay A, Vandame J, Popovici T, Meritet J -F., Cynober L, et al. Diagnostic accuracy of des-gamma-carboxy prothrombin for hepatocellular carcinoma in a French cohort using the Lumipulse ^®^ G600 analyzer. Journal of Viral Hepatitis 2017;24:80–5. 10.1111/jvh.12622.

[34] Tang X-Q, Li H, Yan L-B, Zhou L-Y, Chen E-Q, Liu M, et al. Diagnostic value of PIVKA-II in detecting hepatocellular carcinoma. Future Virology 2017;12:259–67. 10.2217/fvl-2017-0017.

[35] Tran VT, Phan TT, Nguyen TB, Le TT, Tran T-TT, Nguyen A-TT, et al. The diagnostic performance of AFP and PIVKA-II models for non-B non-C hepatocellular carcinoma. BMC Res Notes 2023;16:317. 10.1186/s13104-023-06600-y.

[36] Viggiani V, Palombi S, Gennarini G, D’Ettorre G, De Vito C, Angeloni A, et al. Protein induced by vitamin K absence or antagonist-II (PIVKA-II) specifically increased in Italian hepatocellular carcinoma patients. Scandinavian Journal of Gastroenterology 2016;51:1257–62. 10.1080/00365521.2016.1183705.

[37] Volk ML, Hernandez JC, Su GL, Lok AS, Marrero JA. Risk factors for hepatocellular carcinoma may impair the performance of biomarkers: A comparison of AFP, DCP, and AFP-L31. CBM 2007;3:79–87. 10.3233/CBM-2007-3202.

[38] Wang Q, Chen Q, Zhang X, Lu X-L, Du Q, Zhu T, et al. Diagnostic value of gamma-glutamyltransferase/aspartate aminotransferase ratio, protein induced by vitamin K absence or antagonist II, and alpha-fetoprotein in hepatitis B virus-related hepatocellular carcinoma. WJG 2019;25:5515–29. 10.3748/wjg.v25.i36.5515.

[39] Wu J, Xiang Z, Bai L, He L, Tan L, Hu M, et al. Diagnostic value of serum PIVKA-II levels for BCLC early hepatocellular carcinoma and correlation with HBV DNA. CBM 2018;23:235–42. 10.3233/CBM-181402.

[40] Wu M, Liu Z, Li X, Zhang A, Li N. Dynamic Changes in Serum Markers and Their Utility in the Early Diagnosis of All Stages of Hepatitis B-Associated Hepatocellular Carcinoma. OTT 2020;Volume 13:827–40. 10.2147/OTT.S229835.

[41] Yang T, Xing H, Wang G, Wang N, Liu M, Yan C, et al. A Novel Online Calculator Based on Serum Biomarkers to Detect Hepatocellular Carcinoma among Patients with Hepatitis B. Clinical Chemistry 2019;65:1543–53. 10.1373/clinchem.2019.308965.

[42] Yu R, Xiang X, Tan Z, Zhou Y, Wang H, Deng G. Efficacy of PIVKA-II in prediction and early detection of hepatocellular carcinoma: a nested case-control study in Chinese patients. Sci Rep 2016;6:35050. 10.1038/srep35050.

[43] Li Q, Pan X, Zhu D, Deng Z, Jiang R, Wang X. Circular RNA MAT2B Promotes Glycolysis and Malignancy of Hepatocellular Carcinoma Through the miR-338-3p/PKM2 Axis Under Hypoxic Stress. Hepatology 2019;70:1298–316. 10.1002/hep.30671.

[44] Duval S, Tweedie R. Trim and fill: A simple funnel-plot-based method of testing and adjusting for publication bias in meta-analysis. Biometrics 2000;56:455–63. 10.1111/j.0006-341x.2000.00455.x.

[45] Jia X, Liu J, Gao Y, Huang Y, Du Z. Diagnosis accuracy of serum glypican-3 in patients with hepatocellular carcinoma: a systematic review with meta-analysis. Arch Med Res 2014;45:580–8. 10.1016/j.arcmed.2014.11.002.

[46] Yang J, Li J, Dai W, Wang F, Shen M, Chen K, et al. Golgi protein 73 as a biomarker for hepatocellular carcinoma: A diagnostic meta-analysis. Exp Ther Med 2015;9:1413–20. 10.3892/etm.2015.2231.

[47] Kobeissy A, Merza N, Al-Hillan A, Boujemaa S, Ahmed Z, Nawras M, et al. Protein Induced by Vitamin K Absence or Antagonist-II Versus Alpha-Fetoprotein in the Diagnosis of Hepatocellular Carcinoma: A Systematic Review With Meta-Analysis. J Clin Med Res 2023;15:343–59. 10.14740/jocmr4951.

[48] Hu B, Tian X, Sun J, Meng X. Evaluation of Individual and Combined Applications of Serum Biomarkers for Diagnosis of Hepatocellular Carcinoma: A Meta-Analysis. IJMS 2013;14:23559–80. 10.3390/ijms141223559.

[49] Dai M, Chen X, Liu X, Peng Z, Meng J, Dai S. Diagnostic Value of the Combination of Golgi Protein 73 and Alpha-Fetoprotein in Hepatocellular Carcinoma: A Meta-Analysis. PLoS ONE 2015;10:e0140067. 10.1371/journal.pone.0140067.

[50] Chen H, Chen S, Li S, Chen Z, Zhu X, Dai M, et al. Combining des-gamma-carboxyprothrombin and alpha-fetoprotein for hepatocellular carcinoma diagnosing: an update meta-analysis and validation study. Oncotarget 2017;8:90390–401. 10.18632/oncotarget.20153.

[51] Pang B, Leng Y, Wang X, Wang Y, Jiang L. A meta-analysis and of clinical values of 11 blood biomarkers, such as AFP, DCP, and GP73 for diagnosis of hepatocellular carcinoma. Annals of Medicine 2023;55:42–61. 10.1080/07853890.2022.2153163.

